# Phase I single center trial of Ketogenic Diet for Adults with Traumatic Brain Injury

**DOI:** 10.1101/2021.07.19.21260800

**Authors:** Niraj Arora, N. Scott Litofsky, Golzy Mojgan, Rachna Aneja, Danielle Staudenmyer, Kathyrn Qualls, Sachin Patil

**Affiliations:** University of Missouri, Columbia, Missouri, United States; Molecular Microbiology and Immunology, University of Missouri, Columbia, Missouri, United States; Department of Internal Medicine, University of Missouri, Columbia, Missouri, United States

**Keywords:** Traumatic Brain injury, Ketogenic diet, ketosis, beta-hydroxybutyrate, glucose

## Abstract

**Background:** Traumatic Brain injury (TBI) is a major cause of mortality and morbidity in the United States. Ketogenic diet (KD) has been shown to have neuroprotective effects in acute brain injury, but limited data about its use in adult TBI patients is available. The objective of this study is to investigate the feasibility and safety of ketogenic diet (KD) for adult TBI patients in the Neuroscience Intensive Care Unit (NSICU).

**Methods:** TBI patients admitted to NSICU between June 2019 to March 2021 were enrolled in this single-center, open label, single-arm prospective intervention study. The primary feasibility outcome was achievement of ketosis (detection and maintenance of serum beta-hydroxybutyrate (BOB) levels above normal); secondary outcomes included laboratory and clinical adverse effects related to KD.

**Results:** 10 adults with TBI with Abbreviated Injury Score (AIS)-Head of at least 3 and ventriculostomy tube inserted met inclusion/exclusion criteria and were considered for KD. Mean age was47 years, and all patients were male. Eight out of 10 patients achieved ketosis within mean 2.2 days. KD was initiated within 8-33 hours (average 23 hours) of hospital admission. No clinical adverse effects were noted, 2 patients developed hypertriglyceridemia and 1 patient developed hypoglycemia. Serum glucose showed a decreasing trend in most patients.

**Conclusions:** This pilot study shows that KD is feasible and safe in the management of TBI patients. A randomized controlled trial (RCT) is justified to further understand the optimal serum BOB levels, dose and duration of KD in TBI and its effect on the outcome.

ClinicalTrials.gov Identifier: NCT03982602, Registered 06/11/2019, https://clinicaltrials.gov/ct2/show/NCT03982602?term=brain+injury&cond=ketogenic+diet&draw=2&rank=3.

## Introduction

Traumatic brain injury (TBI) is a major cause of death and disability in young adults contributing to nearly one-third of injury related deaths in the United States. An estimated 13.5 million individuals live with disability after TBI in the United States, creating and significant economic burden on the health-care system and the family[1]. TBI is complicated by development of secondary cerebral injury involving a host of cellular and molecular cascades that promote cell death leading to worsening of cerebral edema and ischemia[2], [3]. Disturbances in ion homeostasis, glutamate excitotoxicity, neuronal depolarization, generation of nitric oxide and oxygen free radicals, lipid peroxidation, and mitochondrial dysfunction results in inflammation, apoptosis, and necrotic cell death [2]. In TBI with initial Glasgow Coma Scale (GCS) 3-5, survival is only 20% and out of that, only 50% patients have good functional outcome. [4] Therefore, development of new strategies and approaches to understand and reverse the neuronal damage is essential. One proposed treatment is the use of a high fat-containing diet known as the Ketogenic Diet (KD).

KD, developed in 1920s, mimics the biochemical environment present during a period of limited food availability [5]. The diet consists of high fat, adequate protein, and very minimal carbohydrates which enhance cellular metabolic and mitochondrial functions [6]. Medium chain fatty-acids derived from the KD have been shown to have a therapeutic role in management of different neurological disorders[5], [7]–[11]. In animal studies with TBI, KD decreases the oxidative stress response and improves mitochondrial function[12]. A corresponding reduction in lactate levels occurs and improvement in energy metabolism occurs in animals treated with KD compared to standard carbohydrate diet [12]. In humans with TBI, administration of glucose leads to inhibition of ketogenesis and increases production of lactate levels in the brain [12]. After TBI, metabolic changes including hyperglycemia, high cerebral glutamate levels, and high lactate/pyruvate ratio occur [2], [3]. These changes lead to poor clinical outcomes after TBI[13]. KD has been tried as a therapy in number of chronic neurological disease like Alzheimer’s Disease (AD)[7], childhood epilepsy[14],[15],and Parkinson’s Disease [16]. While KD has been considered as a safe, feasible and effective therapeutic option in status epilepticus patients due to various causes including TBI, the safety and feasibility of KD in adult humans with TBI has not been studied [10], [17].

The current Phase I study describes the single center ICU experience regarding the safety and feasibility of KD for TBI patients. We hypothesize that KD may be safe and feasible in patients with TBI in ICU.

## Methods

### Standard Protocols and Consents

The study was approved by the University of Missouri Institutional Review Board (IRB), Protocol number 2014675, ClinicalTrials.gov Identifier: NCT03982602. Appropriate consent for the study was obtained from the patient’s next of kin or designated power of attorney at the time of admission. Due diligence was taken to adhere to the ethical standards as defined by the IRB.

### Study design

Adult patients greater than 18 years old admitted to Neuroscience Intensive Care Unit (NSICU) at University of Missouri Hospital and Clinics, Columbia, USA (United States of America) between June 2019 to March 2021 with TBI with Abbreviated Injury Score (AIS)-Head of at least 3 and ventriculostomy catheter or intracranial pressure monitor inserted were eligible to participate in this single-center, open label, single-arm prospective intervention Phase I study to evaluate the safety and feasibility of ketogenic diet in TBI patients. Exclusion criteria[17] included patients with diabetic ketoacidosis, hypoglycemia (glucose <70mg/dl), acute liver failure (liver enzymes x5upper limit of normal and/or total bilirubin > 15mg/dl), hypertriglyceridemia (TG > 200mg/dl), history of pancreatitis, inability to tolerate enteral feeds including ileus, diagnosed case of fatty acid oxidation disorder or pyruvate carboxylase deficiency, pregnant females, death or withdrawal of care within 48 hours of admission, inability to obtain consent from the patient’s decision maker.

### Study Outcomes

Primary feasibility outcome of the study was achievement of ketosis: detection and maintenance of serum beta-hydroxybutyrate (BOB) levels above normal [> 0.27mmol/l][18]. Serum BOB levels were measured before initiating KD (baseline) and then daily. Main safety outcome of the study was development of **laboratory abnormalities** such as hypoglycemia :glucose <70mg/dl, mild, moderate or severe hypertriglyceridemia (Triglyceride{TG} levels 150-199 mg/dL (1.7-2.3 mmol/L), 200-999 mg/dL (2.3 to 11.3 mmol/L), or 1000-1999mg/dL (11.3-22.6 mmol/L respectively) [19], metabolic acidosis (pH<7.2), significant electrolyte disturbances (sodium less than 130mg/dl or more than 150mg/dl, potassium less than 3mg/dl or more than 5mg/dl), acute liver failure (liver enzyme elevation x5 times the upper limit of normal), acute kidney injury (elevation in serum creatinine above normal for age) and **clinical adverse effects** like nausea, vomiting, abdominal distension, diarrhea.

Glucose levels were measured daily while fasting lipid levels were measured weekly. Lowest level of blood glucose during the day was reported while the patient was on KD. The requirement of the insulin per day for each patient was calculated while on KD.

### Patient Management

All patients admitted to the NSICU with severe TBI received standard of care management per the Brain Trauma Foundation guidelines[20]. Surgical evacuation with craniotomy or craniectomy, external ventricular drain (EVD) placement for cerebrospinal fluid (CSF) diversion and intracranial pressure (ICP) monitoring, hypertonic solutions as indicated, sedation, analgesia and paralytics as needed with appropriate medications, antibiotics for infection, seizure prophylaxis, and mechanical ventilation were provided. Marshall Computerized Tomography (CT) score [21] was determined on all included patients. Utmost care was taken to provide the medications that contain minimal or no carbohydrates [22] and the doses were verified by the pharmacist.

### Nutritional management

All patients enrolled in the study protocol were given glucose-free intravenous fluids during resuscitation in emergency room, operating room, or NSICU. Ketogenic diet was started as soon as the patient was ready for alimentation (while they were in NSICU). The rate of feeds was calculated by a trained dietician. Ketogenic diet was continued via nasogastric or orogastric tube during the entire NSICU LOS with inability to swallow. In all other conditions, KD was replaced with the standard carbohydrate diet. The duration of KD therapy was determined and side effects and complications of KD were monitored. If the patient was not able to tolerate the KD due to side effects and/or developed abdominal distension or ileus, then KD was discontinued for 24 hours before resumption. Confounding factors in the form of excessive sedation and medications were managed per the treating intensivist.

Resting Metabolic Rate (RMR) was calculated using the Penn State Formular, as follows: RMR = (Mifflin-St. Jeor x 0.96) + (VE x 32) + (Tmax x 167) – 6212, where VE is minute ventilation (L/min) and Tmax is maximum body temperature in the previous 24 hours in° Celsius. with the Mifflin-St Jeor formula following:

**Men:** (10 × weight in kg) + (6.25 × height in cm) - (5 × age in years) + 5

**Women:** (10 × weight in kg) + (6.25 × height in cm) - (5 × age in years) - 161

Protein requirement (1.5-2 gm/kg of body weight) were met with Prosource ®, which contains only protein and no carbohydrate. If the patient was receiving propofol for sedation, the calories of its lipid content were factored into while calculating the total daily calorie input. Attempt was made to wean propofol off as early as possible to avoid the potential confounding toxic side effects of propofol. Clevedipine infusions, were changed to nicardipine infusions for blood pressure control, since they also contain lipids.

### Descriptive Study Statistics

A sample size of 10 patients was chosen for this phase I pilot study [23]. Proportions were calculated for categorical variables, and means, medians and interquartile ranges (IQRs) were calculated for continuous variables as appropriate. Graphical techniques were used for visualization of trends in glucose, and serum BOB levels with time. A simple regression analysis was used to estimate the daily change in these values. Due to small sample size, we did not include the random variability among the individuals. We looked at the relationship between BOB and time using a simple linear regression.

## Results

### Patient Characteristics

Ten patients identified who met the study inclusion/exclusion criteria (Figure1) were eligible and started on KD therapy. The mean age of included patients was 47.7years with ±15.02 (range 28 – 76 years); all patients were Caucasian males. Only 1 patient had type 2 diabetes mellitus on admission, 4 patients had history of alcohol abuse, 4 patients had history of smoking, 2 patients had methamphetamine abuse, 1 patient had history of hypertension, arrythmias and chronic kidney disease.

**Figure 1:**
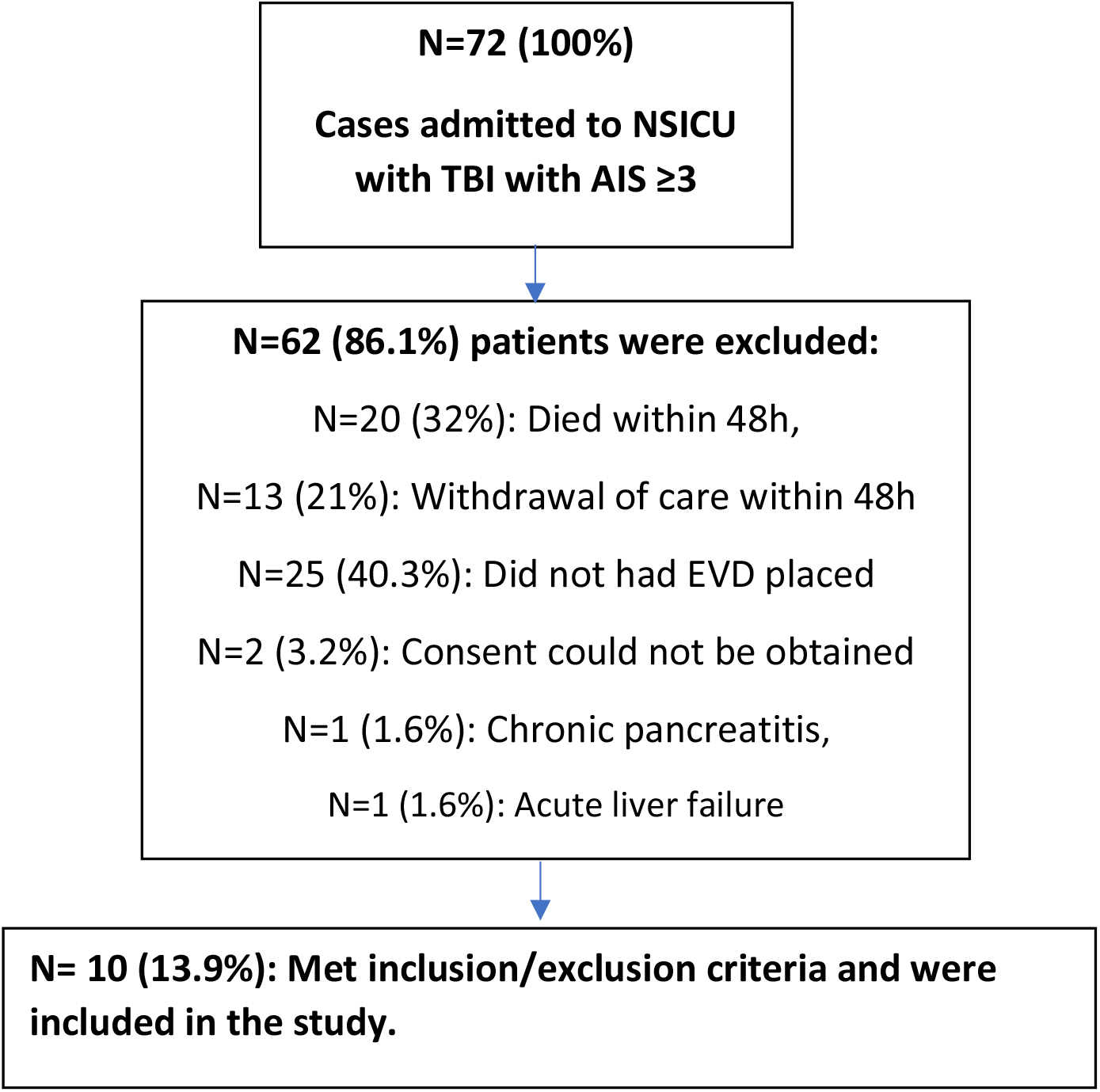
Flowchart screening patients using inclusion/exclusion criteria

Etiology of TBI was related to fall from height (5), fall due to drug intoxication (2), motor vehicle accident (1), mechanical fall (1), and direct trauma to head due to firework mortar (1). Marshall Computerized Tomography (CT) Score was 5 in 7 patients, 4 in 1 patient and 2 in 2 patients. Abbreviated Injury Score (AIS)-Head was 5 in 8 patients, 4 and 3 in 1 patient each. Craniectomy was performed in 2 patients, craniotomy in 5 patients, and all patients had EVD. Eight patients were on propofol infusion for sedation management initially. The maximum dose of the propofol infusion was 55 mcg/kg/hour, which was gradually weaned off once the decision was made to start the patient on KD. Admission GCS was < 8 in all patients except in one. However, patient’s GCS declined to 5t (E1V1tM3) within 24h of ICU admission and prompted urgent surgical intervention.

### Ketogenic diet ratio

All the patients were started on standard formulation of ketogenic diet (Ketovie 4:1) (Table 1). After the correction of the protein requirement, the net ratio averaged 1.33: 1 (range: 1.1-1.76:1).

**Table 1:** Derivation of the fat:Carbohydrate-Protein ratio (F:CP)

| <u>Patient</u> | <u>IBW (kg)</u> | <u>Admission weight (Kg)</u> | <u>Calorie Requirement (Kcal/day)</u> | <u>Protein Requirement (grams/day)</u> | <u>Calories from ketovie 4:1 (Kcal)</u> | <u>Protein from ketovie 4:1 (grams)</u> | <u>Net CHO from ketovie 4:1</u> | <u>Protein supplement (Grams)</u> | <u>Calories from supplement (Kcal)</u> | <u>Fat intake from Ketovie 4:1(grams/day)</u> | <u>Ratio of Fat:Protein+ net CHO</u> |
| --- | --- | --- | --- | --- | --- | --- | --- | --- | --- | --- | --- |
| 1 | 86.4 | 118.8 | 2160 | 132.5 | 1800 | 42.5 | 1.24 | 90 | 360 | 177 | 1.3:1 |
| 2 | 72.7 | 83.6 | 1742 | 122.6 | 1382 | 32.6 | 0.96 | 90 | 360 | 136 | 1.1:1 |
| 3 | 82.3 | 92.8 | 2220 | 147.5 | 1800 | 42.5 | 1.24 | 105 | 420 | 177 | 1.19:1 |
| 4 | 78 | 87 | 2088 | 133 | 1728 | 40.8 | 1.2 | 90 | 360 | 170 | 1.28:1 |
| 5 | 70 | 87.1 | 2088 | 130.8 | 1728 | 40.8 | 1.2 | 90 | 360 | 170 | 1.28:1 |
| 6 | 67.3 | 64.6 | 2140 | 105 | 1900 | 44.9 | 1.32 | 60 | 240 | 187 | 1.76:1 |
| 7 | 86.3 | 135 | 1855 | 112 | 1555 | 36.7 | 1.08 | 75 | 300 | 153 | 1.36:1 |
| 8 | 70 | 127.9 | 1915 | 126.7 | 1555 | 36.7 | 1.08 | 90 | 360 | 153 | 1.35:1 |
| 9 | 78.2 | 66.2 | 1569 | 118.6 | 1209 | 28.6 | 0.82 | 90 | 360 | 119 | 1.1:1 |
| 10 | 64.5 | 53.6 | 1449 | 88.6 | 1209 | 28.6 | 0.84 | 60 | 240 | 119 | 1.33:1 |

IBW: Ideal Body Weight, CHO: carbohydrate

#### Outcomes after KD initiation (Primary outcome)

Eight out of ten patients achieved ketosis and maintained ketosis after initiation of KD (Table 2). The presence of ketones in the serum was seen as early as day 1 (average 2.2 days ± 2.14 days, range 1 - 8 days). The duration of KD ranged from 4 - 14 days, with most patients receiving the therapy for at least 1 week. Four patients had elevated BOB levels before starting of KD which may be related to fasting [24]. KD was initiated within 8-33 hours (average 23 hours) of hospital admission.

**Table 2:** Feasibility, safety and tolerability outcomes of KD.

| Patient | Time for KD initiation from hospital admission (hours) | Baseline BOB level (mmol/l) | Ketosis achieved | Time to ketosis (days) | Laboratory adverse events | Clinical adverse effects | Duration of KD (days) |
| --- | --- | --- | --- | --- | --- | --- | --- |
| 1 | 24 | <0.18 | Yes | 1 | Hyper-Tg (540mg/dl) | No | 9 |
| 2 | 16 | <0.18 | No | - | none | No | 4 |
| 3 | 30 | 0.55 | Yes | 8 | none | No | 14 |
| 4 | 30 | 0.58 | Yes | 3 | none | No | 12 |
| 5 | 12 | 1.53 | Yes | 1 | none | Mild Abdominal pain | 12 |
| 6 | 33 | 0.39 | Yes | 1 | none | No | 6 |
| 7 | 30 | <0.18 | Yes | 1 | none | No | 6 |
| 8 | 30 | 0.23 | No | - | Hyper-Tg (325mg/dl) | No | 10 |
| 9 | 8 | <0.18 | Yes | 1 | none | no | 4 |
| 10 | 18 | 0.29 | yes | 1 | Hypoglycemia | no | 8 |

Hyper-Tg: Hypertriglyceridemia

Two patients did not achieve ketosis during the ICU stay (patient 2, 8). On evaluation, the net KD ratio was 1.1:1 for patient 2. The family opted for withdrawal of care due to refractory ICP elevation. Patient 8 did not achieve ketosis until day 10 when propofol was added to the KD. The net KD ratio was 1.35:1. However, the triglyceride levels increased to 325mg/dl. The patient was continued on KD but he remained in comatose condition throughout the NSICU stay and family decided to discontinue the care. For patient 9, therapy was stopped after 4 days as he recovered neurologically and was extubated and ventriculostomy was removed. However, patient had other medical issues after the extubation which prompted him to stay in the hospital for 15 additional days. The remaining patients achieved ketosis as measured with serum BOB levels above 0.27mmol/l. Patient 7 continued to have poor neurological exam and family decided to withdraw the care. The patient’s serum BOB levels remained low despite being on KD. (Net KD ratio 1.36:1) Serum BOB levels fluctuated for each patient (Figure 2). However, a linear relationship between serum BOB and time was observed (Figure 3).

**Figure 2:**
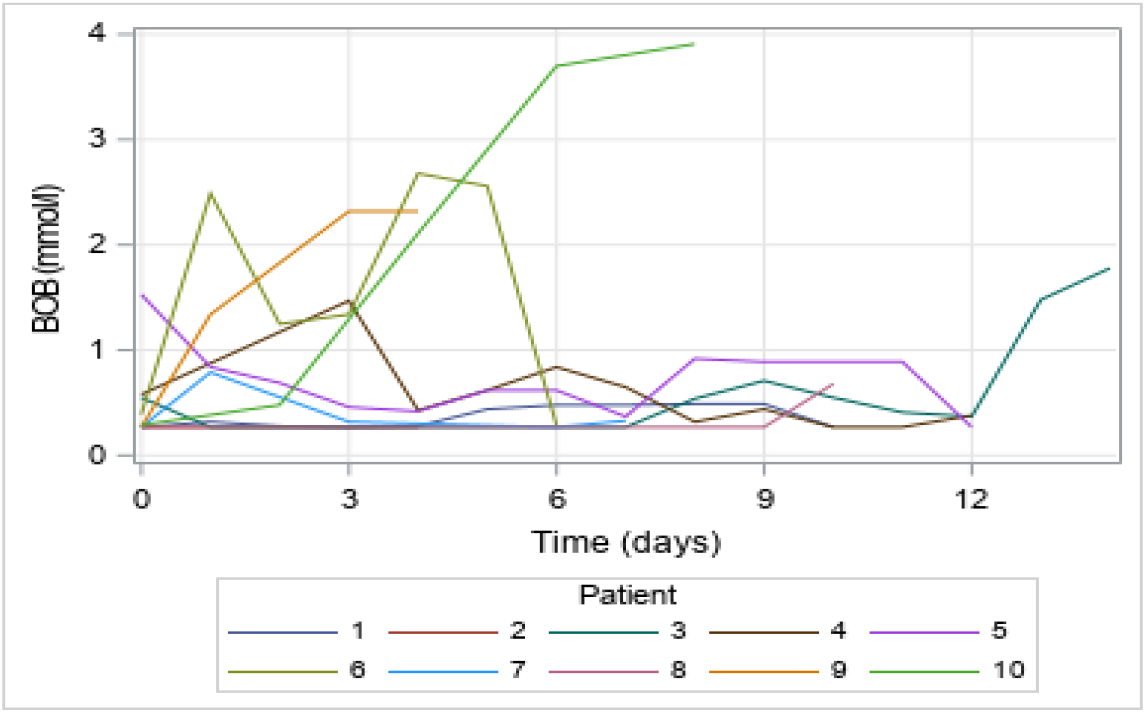
Plots of serum BOB levels (mmol/l) with time for each patient. Note: BOB values <0.27 were truncated to 0.27. There is fluctuation in the values of the serum BOB levels.

**Figure 3:**
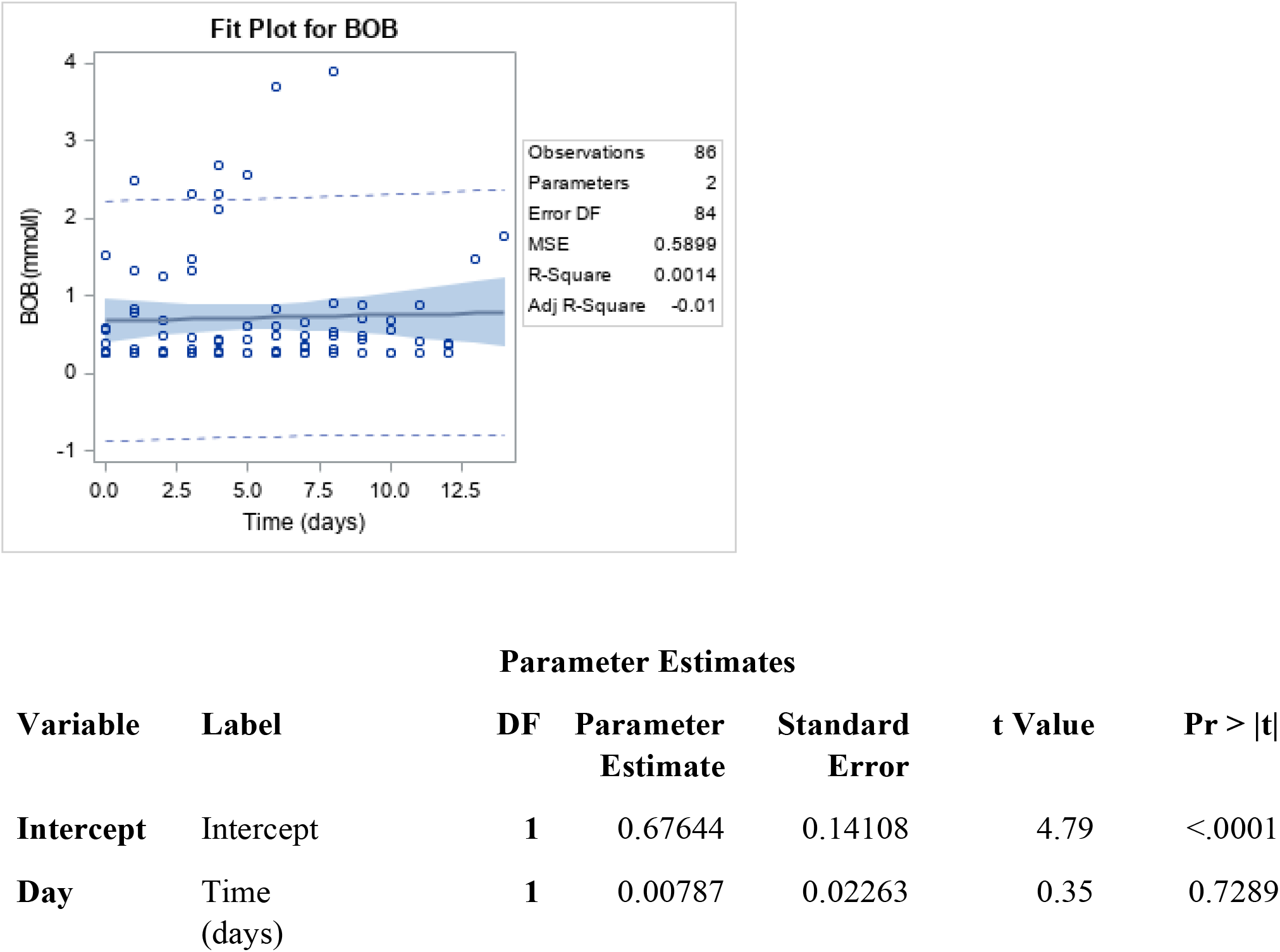
Linear relationship between serum BOB levels (mmol/l) and time (days).

#### Secondary outcomes (Table 3)

No patients had metabolic complications, such as metabolic acidosis, significant electrolyte disturbances, acute liver failure, acute kidney injury or troponinemia. None of the patients developed acute liver failure or propofol infusion syndrome related to the use of propofol with KD.

Two patients (patient 1 and 8) developed hyper-triglyceridemia. Serum blood glucose levels decreased with time in all patients, even if the patient had not achieved a ketotic state. One patient (patient 10) developed hypoglycemia which required dextrose infusion once. The case was adjudicated and it was determined that KD was the cause of hypoglycemia. However, he did not have any recurrent episodes of hypoglycemia and tolerated the KD.

### Effect on glucose and insulin requirement

Glucose levels showed a decreasing trend (Figure 4) and the fitted regression model showed an estimated 3.9mg/dl daily decrease (Figure 5). The individual random variation in glucose level was not considered in the fitted model due to the small number of patients.. After initiation of KD, only 3 patients required insulin. Patient 2 and 9 required 3 units of insulin on day 1 and patient 7 who had poorly controlled diabetes on admission continued to have increased insulin requirement throughout the ICU stay. (Supplementary table 2)

**Figure 4:**
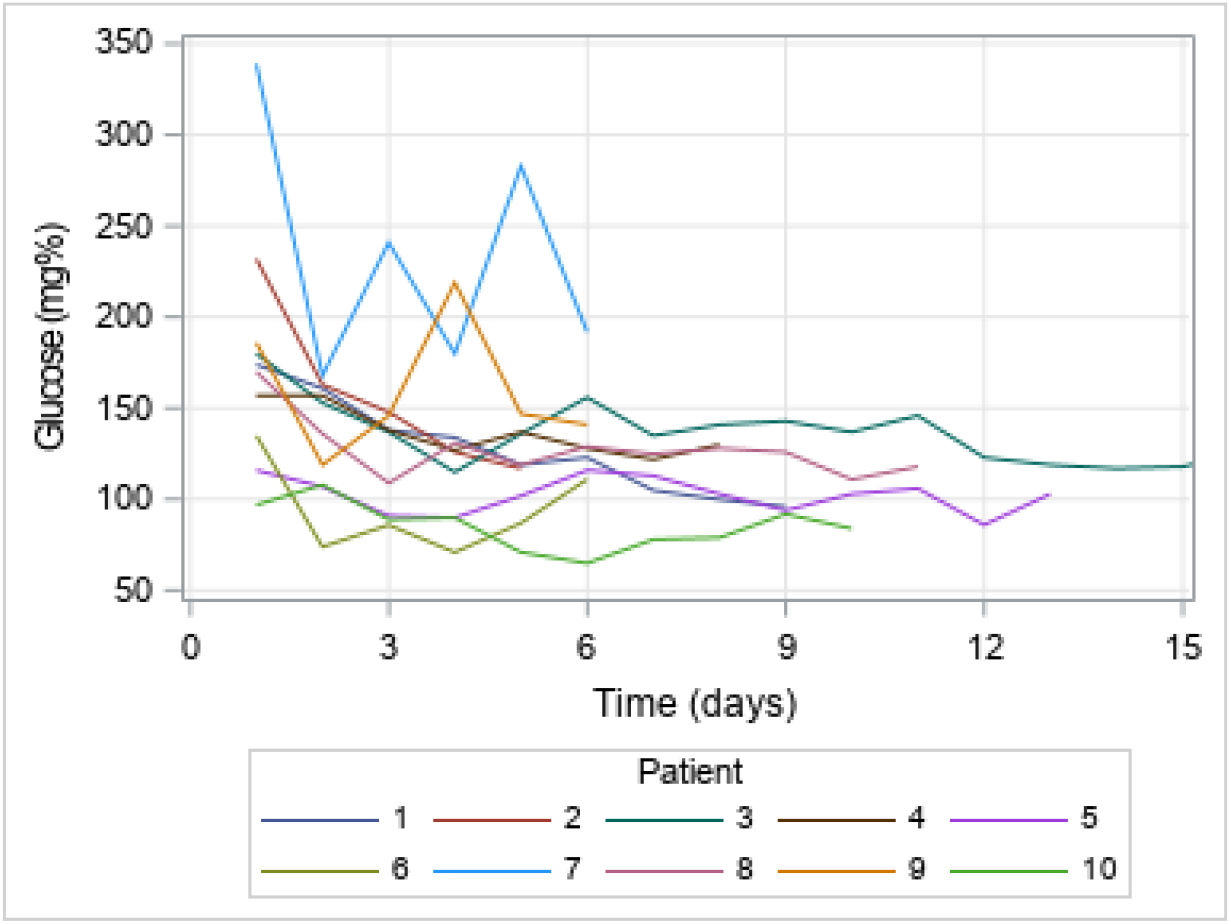
Plot of serum glucose (mg%) with time for each patient. There is decrease in serum glucose levels with time.

**Figure 5: :**
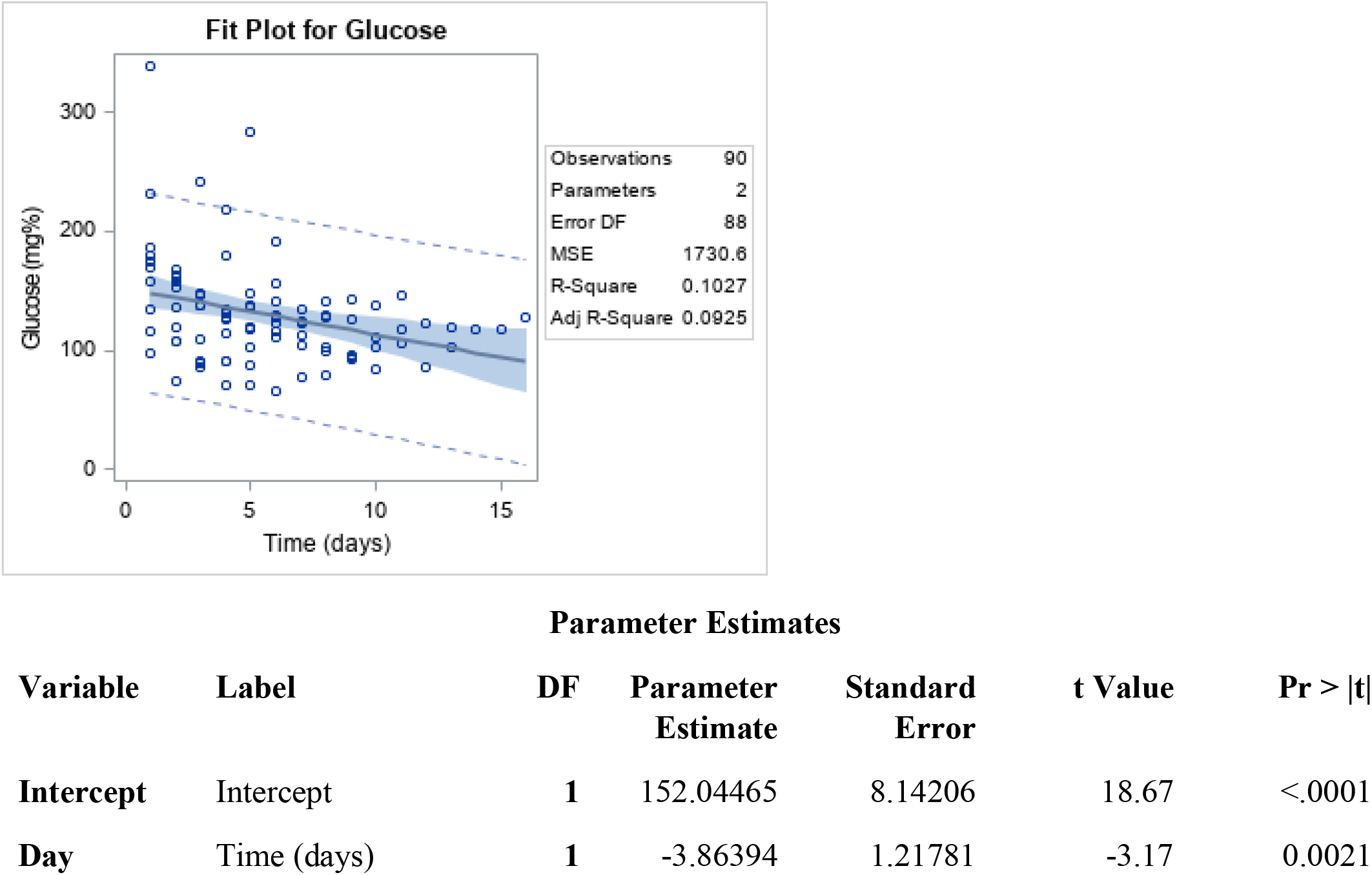
Linear relationship between Glucose levels (mg%) and time (days).

## Discussion

We report a phase I pilot study of TBI patients treated with KD. Most patients achieved ketosis and tolerated the KD without significant clinical adverse effects. Our results about safety of KD are similar to those of patients with status epilepticus. [10], [17], [25]. Adverse effects like hypoglycemia and hypertriglyceridemia were noted in some of the patients. Attempts were not made to achieve higher ketosis levels in our study. Fasting can influence the BOB levels and levels as high as 1mmol/l can be achieved during fasting and starvation[24] however this may not be a feasible and safe option in critical brain injuries. Maintenance of ketosis is essential to understand the beneficial effects of ketones. Serum BOB levels are usually undetectable in patients fed on carbohydrate containing diet.

Previous studies have recommended avoiding propofol when patients are provided KD because of the risk of propofol infusion syndrome[26], [27]. However, use of propofol with KD is probably safe as long as given at low doses (25 – 55 mcg/kg/min) and slowly weaned. The fat component of propofol can affect the ketosis[28], [29] and the calories of patients on propofol need to be adjusted. However, the use of propofol with KD should be tested in larger clinical studies.

Since TBI is a catabolic state[30]–[32], we considered using higher protein content which resulted in lower net fat: carbohydrate plus protein ratio (F:CP). Recent animal studies have found that a ratio of 2:1 formula is better for adults compared to 4:1[33] however that data may not be applicable to adult human TBI patients. In status epilepticus patients, targeting to achieve ketosis with serum BOB levels above 2mmol/l helps to better control seizures [17]. No optimal cut-off for serum BOB levels in TBI patients is available, so we followed the trend of serum BOB levels. Whether providing high fat content only without protein is more beneficial needs to be determined in future studies.

Hyperglycemia has been associated with worse outcome in neurologically injured patients [34]– [36]. The recommended blood glucose levels in the ICU setting are 140 - 180 mg% [36]. Transient hypoglycemia requiring dextrose infusion is uncommon (only 1 of 10 patients). Therefore, the KD may help avoid hyperglycemia without adverse side effects.

### Fat to protein-carbohydrate ratio

In our study we used Ketovie 4:1 formula which provided higher fat, but not enough proteins. Patients require supplemental protein to fulfill the requirement. These supplements, after correction of the calculation with protein, lower the net F:CP ratio. Medium chain triglycerides (MCT) supplementation can be tried to achieve higher ketosis[22] but further studies are needed to understand the effectiveness of higher serum BOB levels.

### Potential basis of KD as a therapy

Based on the existing literature, ketones help in neurological recovery and promotes healing. KD has abundant n-3PUFA which has been shown to mitigate the pathological processes involved in TBI such as mitochondrial dysfunction, apoptotic cell death, glutamate mediated excitotoxicity, and oxidative stress and inflammation [31], [32]. KD also ameliorates the effect of hyper glycemia which has been shown to be deleterious after TBI [37], [38]. Even though upregulation of GLUT1,the main receptor for glucose in the brain, occurs [39], decreases in the glucose levels in the brain as detected by microdialysis studies are present [40], [41]. When KD is used, ketones can enter the Krebs’s cycle bypassing glycolysis which helps to generate more adenosine triphosphate (ATP), the main source of energy [33], [42]. KD stabilizes the lactate-pyruvate ratio which is known to be impaired in the TBI [2], [40]. Unidentified mechanisms of KD are also likely present,which are still to be explored.

To date, adult human population data about the use of KD in TBI patients is very limited, and the optimal duration of KD therapy has not been defined. This study is the first pilot trial of use of KD in TBI patients. Some clinical studies have shown that fasting promotes ketone body concentration in cerebral microdialysis (CMD)[43], which suggests a potential therapeutic option in TBI. Use of intravenous ketone body administration has been shown to improve cerebral metabolism and increase in cerebral blood flow [43], [44]. Our strategy was to provide the benefit of oral KD as an alternative energy source when glucose uptake is diminished in brain.

### Limitations

The study has significant limitations. A small sample size and a single center study may impact the external validity. All the patients in our trial were Caucasian males. This could be related to higher incidence of TBI in males compared to females[45]. While female patients would not be expected to have a different outcome when on KD, this limitation needs to be considered in future designing of the studies. Optimal duration of KD therapy is also not defined and we provided the diet only during NSICU stay. Further studies are needed to study the optimal dose and duration of KD in severe TBI patients. We did not measure the cerebral metabolism through microdialysis catheter to understand the role of the KD on the biochemical parameters. However, KD has been shown to have favorable influence on the cerebral metabolism in experimental studies.[46], [47].

## Conclusion

KD is feasible and safe in adult patients admitted in ICU with TBI where a complex dynamic environment of multiple therapies take place. While further prospective studies are merited, the study could serve as the preliminary source for understanding the optimal serum BOB levels, dose and duration of KD in TBI and its effect on the outcome.

## Data Availability

All the data about the manuscript has been included.

## Acknowledgement

We thank Ajinomoto Cambrooke pharmaceuticals for providing the KD formulation (Ketovie 4:1) for thestudy purpose. However, no direct funding was provided for the study purpose.

