## Supplementary material for "Phase I single center trial of Ketogenic Diet for Adults with Traumatic Brain Injury": Supple Table 1

**Supplementary table 1: Serum Beta-hydroxybutyrate (BOB) levels (mmol/l) according to days**

| Patient | BOB levels | | | | | | | | | | | | | | |
|---|---|---|---|---|---|---|---|---|---|---|---|---|---|---|---|
| | Day 0 (Baseline) | Day 1 | Day 2 | Day 3 | Day 4 | Day 5 | Day 6 | Day 7 | Day 8 | Day 9 | Day 10 | Day 11 | Day 12 | Day 13 | Day 14 |
| 1 | <0.18 | 0.32 | 0.28 | 0.18 | 0.28 | 0.44 | 0.48 | 0.48 | 0.49 | 0.49 | | | | | |
| 2 | <0.18 | 0.18 | <0.18 | <0.18 | <0.18 | | | | | | | | | | |
| 3 | 0.55 | 0.18 | 0.18 | 0.22 | 0.18 | 0.18 | 0.18 | 0.23 | 0.54 | 0.71 | 0.55 | 0.41 | 0.37 | 1.48 | 1.78 |
| 4 | 0.58 | - | - | 1.47 | 0.43 | 0.62 | 0.84 | 0.65 | 0.32 | 0.44 | 0.26 | 0.25 | 0.38 | | |
| 5 | 1.53 | 0.84 | 0.69 | 0.46 | 0.42 | 0.62 | 0.62 | 0.37 | 0.92 | 0.89 | - | 0.89 | 0.26 | | |
| 6 | 0.39 | 2.49 | 1.25 | 1.34 | 2.68 | 2.56 | 0.29 | | | | | | | | |
| 7 | <0.18 | 0.79 | - | 0.32 | - | - | 0.27 | 0.33 | | | | | | | |
| 8 | 0.23 | <0.18 | <0.18 | <0.18 | <0.18 | <0.18 | <0.18 | <0.18 | <0.18 | <0.18 | 0.68 | | | | |
| 9 | <0.18 | 1.34 | - | 2.32 | 2.32 | | | | | | | | | | |
| 10 | 0.29 | - | 0.48 | - | 2.11 | - | 3.7 | - | 3.91 | | | | | | |
