## Supplementary material for "Phase I single center trial of Ketogenic Diet for Adults with Traumatic Brain Injury": Supple table 2

**Supplementary table 2: Glucose (mg/dl) trend during ICU stay**

| Patient | 1 | 2 | 3 | 4 | 5 | 6 | 7 | 8 | 9 | 10 |
|---|---|---|---|---|---|---|---|---|---|---|
| ucose on day | | | | | | | | | | |
| 0 | 174 | 232 | 180 | 157 | 116 | 135 | 339 | 170 | 186 | 97 |
| 1 | 161 | 163 | 153 | 157 | 107 | 74 | 168 | 136 | 119 | 108 |
| 2 | 138 | 148 | 137 | 138 | 91 | 86 | 241 | 109 | 146 | 89 |
| 3 | 134 | 126 | 115 | 127 | 90 | 71 | 180 | 131 | 219 | 90 |
| 4 | 119 | 117 | 136 | 137 | 102 | 87 | 243 | 119 | 147 | 71 |
| 5 | 123 | | 156 | 122 | 116 | | 192 | 129 | 141 | 65 |
| 6 | 105 | | 135 | 130 | 113 | | 258 | 125 | | 78 |
| 7 | 100 | | 141 | | 103 | | 193 | 128 | | 79 |
| 8 | 96 | | 143 | | 94 | | 178 | 126 | | 92 |
| 9 | | | 137 | | 103 | | | 111 | | 84 |
| 10 | | | 146 | | 106 | | | 118 | | |
| 11 | | | 123 | | 86 | | | 100 | | |
| 12 | | | 119 | | 103 | | | 111 | | |
| 13 | | | 117 | | | | | | | |
| 14 | | | 118 | | | | | | | |
| 15 | | | 127 | | | | | | | |
