## Supplementary material for "Phase I single center trial of Ketogenic Diet for Adults with Traumatic Brain Injury": Supple table 3

**Supplementary table 3: Insulin requirments for all patients**

| **Patient** | **1** | **2** | **3** | **4** | **5** | **6** | **7** | **8** | **9** | **10** |
|---|---|---|---|---|---|---|---|---|---|---|
| **Regular Insulin (Units)** | | | | | | | | | | |
| Day 0 | none | 3 | none | none | none | none | 66 | none | 3 | None |
| 1 | | | | | | | 50 | | | |
| 2 | | | | | | | 50 | | | |
| 3 | | | | | | | 47 | | | |
| 4 | | | | | | | 47 | | | |
| 5 | | | | | | | 42 | | | |
| 6 | | | | | | | 67 | | | |
| 7 | | | | | | | 61 | | | |
| 8 | | | | | | | 68 | | | |
